# Increasing Postpartum Primary Care Engagement through Default Scheduling and Tailored Messaging*: A Randomized Clinical Trial*

**DOI:** 10.1101/2024.01.21.24301585

**Authors:** Mark A. Clapp, Alaka Ray, Pichliya Liang, Kaitlyn E. James, Ishani Ganguli, Jessica Cohen

## Abstract

**Importance:** Over 30% of pregnant people have at least one chronic medical condition, and nearly 20% develop gestational diabetes or pregnancy-related hypertension, increasing the risk of future chronic disease. While these individuals are often monitored closely during pregnancy, they face significant barriers when transitioning to primary care following delivery, due in part to a lack of health care support for this transition.

**Objective:** To evaluate the impact of an intervention designed to improve postpartum primary care engagement by reducing patient administrative burden and information gaps.

**Design:** Individual-level randomized controlled trial conducted from November 3, 2022 to October 11, 2023.

**Setting:** One hospital-based and five community-based outpatient obstetric clinics affiliated with a large academic medical center.

**Participants:** Participants included English- and Spanish-speaking pregnant or recently postpartum adults with obesity, anxiety, depression, diabetes mellitus, chronic hypertension, gestational diabetes, or pregnancy-related hypertension, and a primary care practitioner (PCP) listed in their electronic health record (EHR).

**Intervention:** A behavioral economics-informed intervention bundle, including default scheduling of postpartum PCP appointments and tailored messages.

**Main Outcome:** Completion of a PCP visit for routine or chronic condition care within 4 months of delivery.

**Results:** 360 patients were randomized (Control: N=176, Intervention: N=184). Individuals had mean (SD) age 34.1 (4.9) years and median gestational age of 36.3 weeks (interquartile range (IQR) 34.0-38.6 weeks) at enrollment. The distribution of self-reported races was 7.4% Asian, 6.8% Black, 15.0% multiple races or “Other,” and 68.6% White. Most (75.8%) participants had anxiety or depression, 15.9% had a chronic or pregnancy-related hypertensive disorder, 19.8% had pre-existing or gestational diabetes, and 40.4% had a pre-pregnancy BMI ≥30 kg/m^2^. Medicaid was the primary payer for 21.9% of patients. PCP visit completion within 4 months occurred in 22.0% in the control group and 40.0% in the intervention group. In regression models accounting for randomization strata, the intervention increased PCP visit completion by 18.7 percentage points (95%CI 10.7-29.1). Intervention participants also had fewer postpartum readmissions (1.7 vs. 5.8%) and increased receipt of the following services by a PCP: blood pressure screening (42.8 vs. 28.3%), weight assessment (42.8 vs. 27.7%), and depression screening (32.8 vs. 16.8%).

**Conclusions and Relevance:** In this randomized trial of pregnant individuals with or at risk for chronic health conditions, default PCP visit scheduling, tailored messages, and reminders substantially improved postpartum primary care engagement. The current lack of support for postpartum transitions to primary care is a missed opportunity to improve recently pregnant individual’s short- and long-term health. Reducing patient administrative burdens may represent relatively low-resource, high-impact approaches to improving postpartum health and wellbeing.

**Trial Registration:** NCT05543265.

**KEY POINTS:** *Question:* What is the impact of a behavioral economic intervention designed to reduce patient administrative burden and information gaps on primary care practitioner (PCP) visit completion in the postpartum period?

*Findings:* In this randomized clinical trial of 360 patients, default PCP scheduling, tailored messages, and reminders increased postpartum PCP visit rates by 19 percentage points for individuals with or at high risk for chronic disease.

*Meaning:* A multi-faceted and relatively low-resource behavioral economic intervention may improve postpartum health and well-being.

## INTRODUCTION

Although the chronic disease burden in pregnancy is high and growing in the U.S., most people with chronic conditions do not effectively transition to primary care management following delivery.^1–9^ Over 30% of pregnant people have diabetes, hypertension, or obesity, and 11-22% have anxiety or depression.^10–12^ Furthermore, common pregnancy-related conditions (e.g., gestational diabetes and pregnancy-related hypertension) confer an increased risk of developing chronic disease.^13–18^ Strong evidence underpins the benefits of managing chronic conditions through primary care and of managing these conditions earlier in life.^19–22^ However, while pregnant people with these conditions are often carefully monitored during pregnancy, many receive no routine care after their pregnancy, and nearly half of those with chronic conditions do not see their primary care practitioner (PCP) at all in the postpartum year.^23^ The abrupt drop off from high health system engagement and motivation during pregnancy to limited or no health care encounters postpartum has been termed a “postpartum cliff.”^24^ Low rates of postpartum primary care engagement reflect a missed opportunity to improve the prevention and management of chronic disease.

Postpartum transitions from obstetric to primary care are encouraged by guidelines yet stymied by numerous barriers. Specifically, obstetric clinical guidelines recommend that all individuals have a comprehensive postpartum visit within 12 weeks of their delivery; at that time, obstetric care clinicians typically counsel patients on the importance of ongoing primary care follow-up. Yet, a range of systemic, financial, and behavioral barriers often prevent postpartum people from effectively transitioning to primary care.^25–29^ Patient administrative burden (e.g., appointment scheduling, information seeking, insurance/billing issues) is increasingly recognized as a barrier to accessing care.^30^ In a recent survey, 33% of patients reported that they delayed or did not seek health care because of the administrative burden.^30^ The effects of this burden may be amplified in the postpartum period when new parents are sleep-deprived and face many competing demands, including caring for their newborn and family. This study aimed to increase patient engagement in primary care after the immediate postpartum period for pregnant individuals with conditions that convey a long-term health risk by reducing administrative burden and motivating continued health activation through an intervention based on insights from behavioral economics.

## METHODS

### Study Design

This study was an individual-level, two-group, 1:1 stratified randomized controlled trial of the effectiveness of a behavioral economics-informed intervention to increase the rate of postpartum primary care visit completion. The study was registered on ClinicalTrials.gov (NCT05543265) on September 6, 2022, and conducted from November 3, 2022, to October 11, 2023 (trial protocol and analysis plan included in Online Supplement). The Mass General Brigham Human Subjects Committee approved this study; participants provided verbal consent to participate. The Consolidated Standards of Reporting Trials (CONSORT) guidelines were followed in reporting the study and its results.

### Patient Eligibility

Patients who had obesity (pre-pregnancy body mass index (≥30 kg/m^2^)), anxiety or depressive mood disorder, type 1 or 2 diabetes mellitus, chronic hypertension, gestational diabetes, or pregnancy-related hypertension listed in their medical record were eligible to participate. Patients at high risk for hypertensive disorders of pregnancy, defined as those who would be recommended for low-dose aspirin by US Preventative Services Task Force guidelines, were eligible. Patients with these conditions were prioritized for inclusion in the study as they were more likely to have ongoing care needs after pregnancy. Also, this study was limited to patients who had a PCP listed or identified in their medical record, as the barriers and solutions to post-delivery primary care re-engagement are different than establishing care with a new PCP; a preliminary analysis of patients receiving obstetric care at the study institution revealed that 90% had a PCP listed in the EHR. Other eligibility criteria included: 1) pregnant or recently postpartum (defined as up to two weeks after their estimated due date (EDD)), 2) receipt of prenatal care at the study institution or its affiliated clinics, 3) enrolled and elected to receive messages in the study institution’s EHR patient portal, 4) primary language of English or Spanish, 5) age ≥18 years at the time of enrollment, and 6) not actively undergoing a work-up for, or known to have, fetal demise at the time of enrollment.

### Enrollment and Randomization

Eligible patients were approached in person and via telephone during the eligibility window (up to two weeks after their EDD). Those who consented to participate in the study were also asked to consent to receive text (SMS) messages separately. Individuals were randomized using a randomization table created by the statistician (KEJ) and uploaded directly into the REDCap randomization module, which was blinded to the primary investigators and study staff. The assignment sequence was stratified by two variables that were determined a priori to be important to ensure balance: 1) visit with a PCP within 3 years before the EDD and 2) site of prenatal care (hospital campus vs. community-based obstetric clinic). Patients were randomized after they consented and completed a baseline survey.

### Study Intervention

The intervention was designed to increase the rate of postpartum primary care visit completion within 4 months after the patient’s EDD. The bundle included a targeted introduction message about the importance of seeing their PCP after delivery and informed them that, to support them in this, a study staff member would be making an appointment on their behalf; they were allowed to opt-out or communicate about scheduling preferences. For those who did not opt-out, the study staff called the PCP office and requested that “health care maintenance” or “annual visit” be scheduled within the target 4-month window. If a patient had already seen their PCP for an annual visit within the year, they were scheduled for this visit when they were next eligible (i.e., one year after their last annual exam), even if outside the 4-month study follow-up period. For those who had appointments scheduled, study-specific appointment reminders were sent approximately 1 month after the EDD and 1 week before the scheduled appointment via the EHR patient portal and SMS, both used salient labeling to describe the visit; examples are shown in Appendix (eFigure 1). If the PCP worked in the same health system and an appointment was scheduled, an electronic message was sent to the PCP from the study staff about the appointment scheduled by the study staff. For those for whom an appointment could not be scheduled, similar reminders were sent on the importance of PCP follow-up and encouraged the patient to contact their PCP’s office directly to schedule. Reminders included best practice wording from behavioral economic nudge “mega-studies,” including that the appointment had been “reserved for you.”^31^ Using salient labeling, the appointment was described as the “Postpartum-to-Primary Care Transition Appointment.”

**Figure 1.**
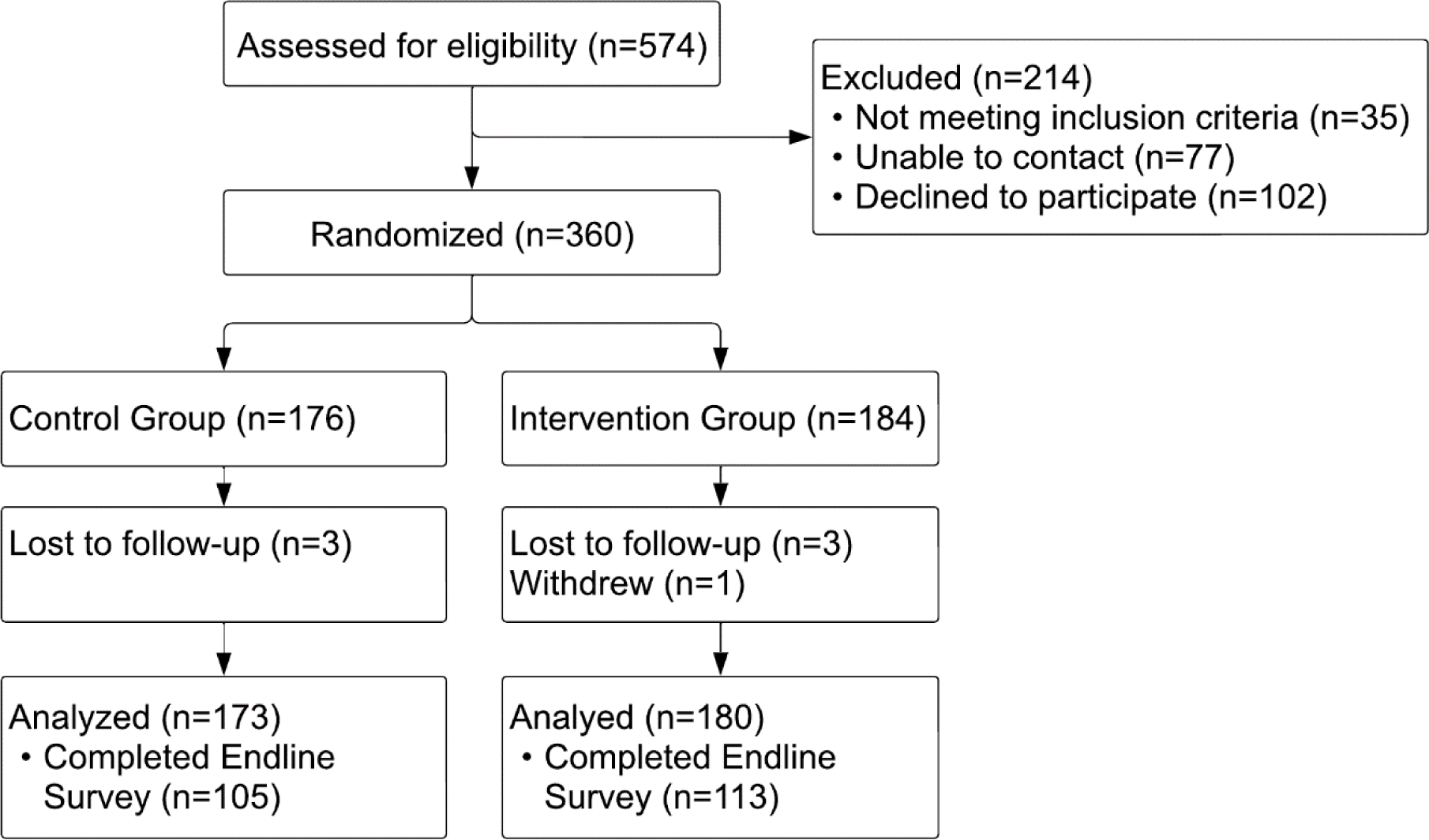
Consort Diagram.

Patients in the control group received 1 message within 2 weeks of the EDD with a generic recommendation for PCP follow-up after delivery.

### Study Measures

The primary outcome was completing a primary care visit for routine or chronic condition care within 4 months of the patient’s EDD. Specifically, we considered the outcome to have occurred if the patient attended a “health care maintenance” (i.e., “annual exam”) visit or a “problem-based visit” in which obesity, anxiety/depression, diabetes, or hypertension were addressed with a primary care clinician within 4 months after their EDD. This definition was chosen to include visits most likely to reflect primary care re-engagement after delivery instead of a visit for an acute illness or issue. This time frame was selected for two reasons: 1) to capitalize on the increased health activation and motivation that has been noted during pregnancy and 2) because these patients were more likely to have conditions that required ongoing and active management outside of the traditional postpartum period (up to 12 weeks after delivery). We considered practitioners affiliated with internal medicine, family medicine, pediatric/adolescent medicine, and gynecology practices to provide primary care; however, we did not count designated postpartum visits to be primary care visits.

Alternate specifications for the primary outcome were compared in sensitivity analyses: 1) self-reported PCP visits within 4 months after the EDD, obtained from a survey sent approximately 5 months after the EDD; 2) primary outcome restricted to visits with the patient’s designated PCP; 3) primary outcome restricted to patients whose PCP was affiliated with the study institution’s health system, 4) primary outcome expanded to include any PCP visit (not only routine or chronic condition care) within 4 months after a patient’s EDD, and 5) primary outcome expanded to include any completed or scheduled PCP visit within 1 year of a patient’s EDD.

We examined secondary outcomes measuring unscheduled care: obstetric triage visit, emergency room or urgent care use, and readmission within 4 months after the delivery. We also measured the likelihood of a patient having a PCP visit that included specific primary care services within 4 months: weight screening, blood pressure screening, mood screening, plan for diabetes screening, plan for mental health care, and contraception planning. Content of care outcomes were also compared within population subgroups related to the eligibility health condition. All outcomes are defined in detail in Appendix (eTable 1).

The primary and most secondary outcomes were ascertained directly by reviewing the patient’s medical record approximately 5 months after their EDD. Study staff that performed the chart review were blinded to the group assignment. Secondary self-reported outcomes were obtained by an electronic survey sent to patients approximately 5 months after their EDD.

### Sample Size Calculation

Based on a historical cohort, we estimated that 33% of the targeted study population would have a PCP visit within 4 months of delivery. We estimated the intervention would increase the rate of PCP visit attendance by at least 15 percentage points, a conservative estimate based on a prior study that examined the impact of default scheduling of postpartum obstetric care appointments (24 percentage point increase).^32^ Assuming an alpha of 0.05 and power of 80% and using a two-sided z-test, 334 patients were needed to detect a 15-percentage point difference. To account for individuals who may be lost to follow-up or withdraw, we planned to randomize 360 patients.

### Statistical Analysis

Patients were analyzed by intention-to-treat. Patients who were lost to follow-up (i.e., transferred obstetric care before delivery) or withdrew before the outcome assessment were excluded. Baseline patient characteristics and the percentage of patients who accessed the study messages in the EHR patient portal were reported. Primary and secondary outcomes were compared using chi-squared, t-tests, and Fisher’s exact test, where appropriate. The percentage point difference in outcomes between the two groups was estimated using a linear probability regression model that included two indicator terms for the randomization strata, which were defined *a priori*.

A heterogeneity analysis was performed to understand the potential impact of the intervention among patient factors known or hypothesized to be disproportionately affected by administrative burdens. The primary outcome was compared among subgroups based on site of prenatal care (hospital-vs. community-based clinic), chronic conditions (anxiety/depression, diabetes, hypertension, obesity, and multi-morbidity, defined as >1 of the listed conditions), race (self-described Asian, Black, White, Other or multiple races), ethnicity (self-described Hispanic or non-Hispanic), individual earnings/income (<$30,000, $30-75,000, or >$75,000), primary payer for delivery hospitalization (Medicaid or Private/Other), and self-reported physical and mental health status at time of enrollment.^30,33^

Stata 16.1 (StataCorp, College Station, TX) was used for the analysis. P-values were reported for the primary outcome; a p-value <0.05 was considered statistically significant. As this project was not designed to have statistical power to detect the intervention’s impact on secondary outcomes or differences across subgroups, a plan for multiple hypothesis testing was not planned or prespecified. Results from secondary analyses are presented with 95% confidence intervals that are not adjusted for multiple hypothesis testing. These secondary analyses should be considered exploratory, and results may not be reproducible.

## RESULTS

Initially, 574 patients were identified as likely to be eligible based on pre-defined eligibility filters within the EHR (Figure 1). Upon chart review, 35 were determined ineligible. Of those confirmed eligible, 77 could not be contacted and 102 declined. 360 patients were randomized: 176 to the control group and 184 to the intervention group. Six patients were excluded from the final analysis because they transferred their care to another institution before delivery (3 in each group). One patient in the intervention group withdrew from the study before the end of the follow-up period. The final number of patients analyzed in each group was 173 in the control group and 180 in the intervention group. Among study participants, 345/353 (97.7%) accessed study-related messages in the online patient portal. The proportion of patients in the intervention group who received each component of the intervention bundle is included in the Appendix (eTable 2); the study staff scheduled appointments for 137 participants (76.1%), of whom only 6 (4.4%) did not present for or cancel their appointment. The most common reason the study staff did not schedule an appointment was that a PCP appointment was already scheduled 21/43 (49%) (see eTable 3 for the full list). Of all participants, 61.8% completed the online electronic survey 5 months after the EDD.

The intervention and control groups were balanced in all baseline patient characteristics (Table 1). Individuals included in the trial had mean (SD) age of 34.1 (4.9) years old and median gestational age of 36.3 weeks (interquartile range (IQR) 34-38.6 weeks) at enrollment. The distribution of self-reported races was 7.4% Asian, 6.8% Black, 15.0% multiple races or “Other,” and 68.6% White; 2.3% declined to report their race. The distribution of self-reported ethnicities was 22.1% Hispanic ethnicity and 75.4% non-Hispanic ethnicity; 2.5% declined to report their ethnicity. Of the eligibility conditions, which were not mutually exclusive, 75.8% of all participants had anxiety or depression, 15.9% had a chronic or pregnancy-related hypertensive disorder, 19.8% had pre-existing or gestational diabetes, and 40.4% had a pre-pregnancy BMI ≥30 kg/m^2^. Medicaid was the primary payer for the delivery encounter for 21.9% of patients. When surveyed, 11.6% reported their physical health and 19.6% reported their mental health as “fair” or “poor” condition. At enrollment, 34.3% had not seen any PCP within the prior 3 years, and 29.2% were receiving obstetric care at one of the hospital’s satellite or affiliated health center clinics.

**Table 1.** Baseline Characteristics of the Analytical Sample.

| Characteristic | Control Group<br>(n=173) | Intervention<br>Group<br>(n=180) |
| --- | --- | --- |
| Patient Age at Estimated Due Date, Mean (SD), Years | 34.0 (5.0) | 34.2 (4.8) |
| Enrollment Relative to Delivery |  |  |
| During Pregnancy | 170 (98.3) | 173 (96.1) |
| Postpartum | 8 (1.7) | 7 (3.9) |
| Gestational Age at Enrollment, Mean (SD), Days | 254 (20) | 255 (20) |
| Primary Site of Prenatal Care |  |  |
| Hospital-based Clinic | 121 (69.9) | 129 (71.7) |
| Community-based Clinic | 52 (30.1) | 51 (28.3) |
| PCP Visit within 3 Years Prior to Enrollment | 121 (69.9) | 111 (61.7) |
| Health Condition |  |  |
| Anxiety or Depression | 128 (74.0) | 138 (76.7) |
| Chronic or Gestational Hypertensive Disorder | 26 (15.0) | 31 (17.2) |
| Chronic or Gestational Diabetes Mellitus | 38 (22.0) | 31 (17.2) |
| Obesity (Pre-pregnancy BMI ≥ 30 kg/m<sup>2</sup>) | 75 (43.4) | 69 (38.3) |
| Race<sup>a</sup> |  |  |
| Asian | 13 (7.5) | 11 (6.1) |
| Black | 12 (6.9) | 14 (7.8) |
| Multiple Races or "Other"<sup>b</sup> | 28 (16.2) | 25 (13.9) |
| White | 115 (66.5) | 127 (70.6) |
| Declined / Not reported | 5 (2.9) | 3 (1.7) |
| Ethnicity<sup>a</sup> |  |  |
| Hispanic | 41 (23.7) | 37 (20.6) |
| Non-Hispanic | 127 (73.4) | 139 (77.2) |
| Not reported | 5 (2.9) | 4 (2.2) |
| Preferred Language<sup>a</sup> |  |  |
| English | 161 (93.1) | 167 (92.8) |
| Spanish | 12 (6.9) | 13 (7.2) |
| Marital Status<sup>a</sup> |  |  |
| Married | 125 (72.3) | 137 (76.1) |
| Not married | 48 (27.7) | 43 (23.8) |
| Education<sup>a</sup> |  |  |
| High School Graduate or Some High School | 30 (17.3) | 22 (12.2) |
| Some College | 11 (6.4) | 22 (12.2) |
| Bachelor's or Associate Degree | 69 (39.9) | 71 (39.4) |
| Graduate School Degree | 63 (36.4) | 65 (36.1) |
| Individual Annual Earnings<sup>a</sup> |  |  |
| <\$30,000 | 32 (18.5) | 36 (20.0) |
| \$30,000 - \$74,999 | 37 (21.4) | 55 (30.6) |
| ≥\$75,000 | 82 (47.4) | 76 (42.2) |
| Not reported | 22 (12.7) | 13 (7.2) |
| Primary Payer for Delivery Hospitalization |  |  |
| Medicaid | 40 (23.1) | 35 (19.4) |
| Private / Other | 130 (75.1) | 138 (76.7) |
| Unknown | 3 (1.7) | 7 (3.9) |
| Physical Health Status at Time of Enrollment<sup>a</sup> |  |  |
| "Good", "Very Good," or "Excellent" | 151 (87.3) | 161 (89.4) |
| "Fair" or "Poor" | 22 (12.7) | 19 (10.6) |
| Mental Health Status at Time of Enrollment<sup>a</sup> |  |  |
| "Good", "Very Good," or "Excellent" | 136 (78.6) | 148 (82.2) |
| "Fair" or "Poor" | 37 (21.4) | 32 (17.8) |

<sup>a</sup> Self-reported.
<sup>b</sup> Patients could select "Other" as a race option if they did not self-identify with the other options: American Indian or Alaskan Native, Asian, Black, Native Hawaiian or Other Pacific Islander, White.
Abbreviation: PCP, Primary care practitioner

Table 2 shows the effects of the intervention on completion of a primary care visit for routine or chronic condition care within 4 months of the patient’s EDD. This primary outcome occurred in 40.0% (95% confidence interval (CI) 33.1-47.4%) of the intervention group and 22.0% (95%CI 6.4-28.8%) of the control group (p<0.001). When adjusting for pre-specified randomization strata, the intervention increased the primary outcome by 18.7 percentage points (pp) (95%CI 9.1-28.2 pp). The effects on the primary outcome were similar in the sensitivity analyses (Table 3). There were no effects on obstetric triage visits or emergency or urgent care use. However, the intervention group had fewer postpartum readmissions: 1.7% (95%CI 0.5-5.1%) vs. 5.8% (95%CI 3.1-10.4%).

**Table 2.**
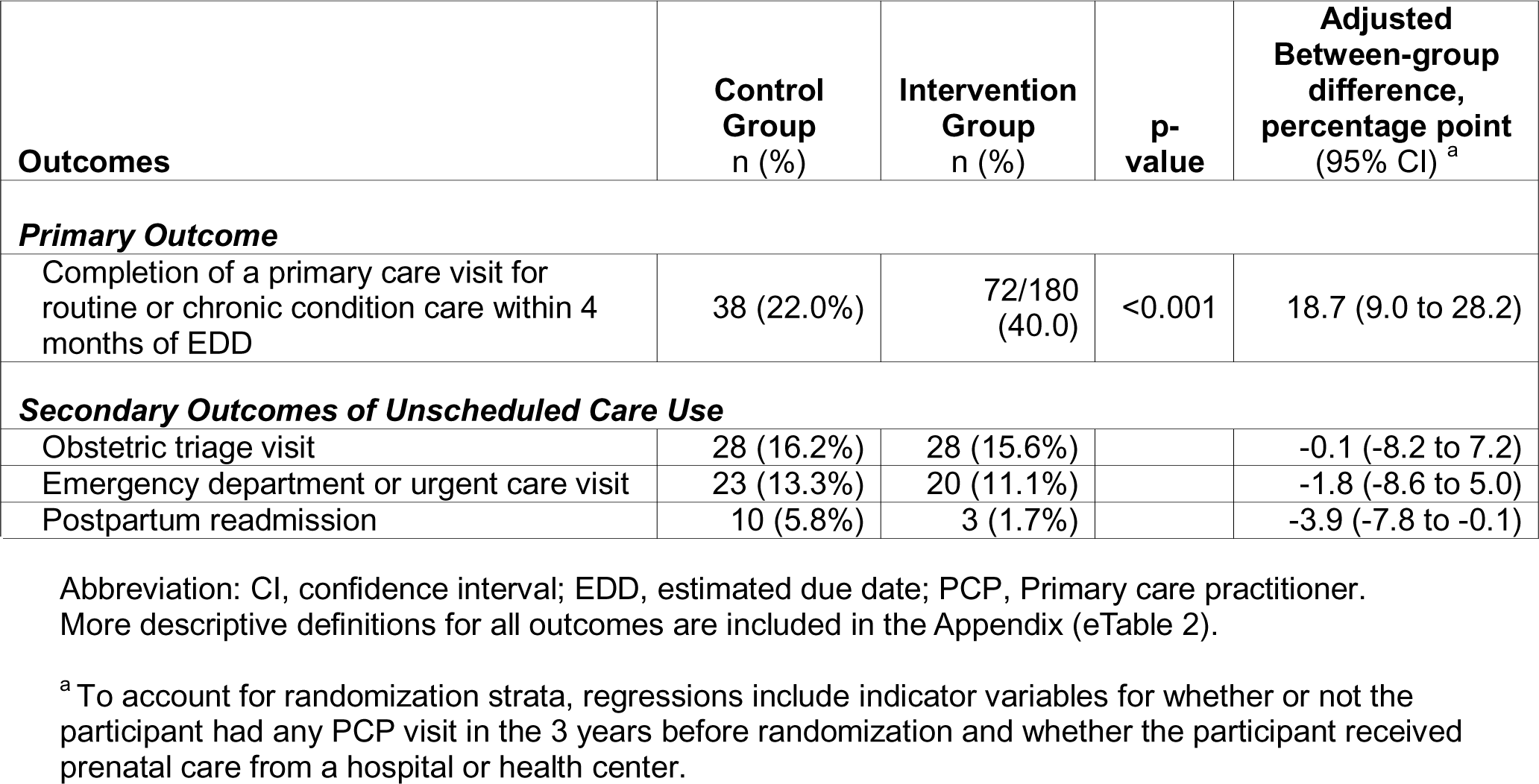
Effects on Care Utilization.

**Table 3.** Sensitivity Analyses for the Primary Outcome.

| **Sensitivity Analysis** | **Control Group**<br>**n (%)** | **Intervention Group**<br>**n (%)** | **Adjusted Between-group difference, percentage point (95 CI)<sup>a</sup>** |
| --- | --- | --- | --- |
| Self-reported PCP visit completion within 4 months of delivery | 46/113 (40.7) | 65/105 (61.9) | 20.1 (7.0 to 33.3) |
| Primary outcome restricted to visits with the patient's designated PCP | 23/173 (13.3) | 50/180 (27.8) | 15.1 (6.7 to 23.5) |
| Primary outcome restricted to patients whose PCP in the same health system | 30/123 (24.4) | 58/118 (49.2) | 24.6 (12.5 to 36.6) |
| Any primary care visit within 4 months of EDD | 54/173 (31.1) | 82/180 (25.6) | 15.1 (5.0 to 25.3) |
| Any primary care visit or scheduled within 1 year of EDD | 75/173 (43.4) | 117/180 (65.0) | 24.0 (14.2 to 33.8) |

<sup>a</sup> To account for randomization strata, regressions include indicator variables for whether or not the participant had any PCP visit in the 3 years before randomization and whether the participant received prenatal care from a hospital or health center.

Figure 2 compares the secondary outcomes related to the content or provision of care between the two groups. Intervention group participants had higher likelihood of having a PCP visit with a weight screening (42.8 (95%CI 35.7-50.1 %) vs. 27.7% (95%CI 21.6-34.9%)), blood pressure screening (42.8 (95%CI 35.7-50.1%) vs. 28.3% (95%CI 22.1-35.1%)), and mood screening (32.8 (95%CI 26.3-40.0%) vs. 16.8% (95%CI 11.9-23.1%)). Intervention group participants were also more likely to have a PCP visit with a plan documented about their mental health (37.2 (95%CI 30.5-44.5%) vs. 23.1% (95%CI 17.4-30.0%)) and with a documented contraception plan (19.4 (95%CI 14.3-25.9%) vs 11.0% (95%CI 7.1-16.6%)). There was no difference in a documented plan for diabetes screening between the two groups. Comparisons of the secondary outcomes related to the content of care among subgroups of health conditions are shown in eTable 4 in the Appendix; many were limited by small sample sizes.

**Figure 2.**
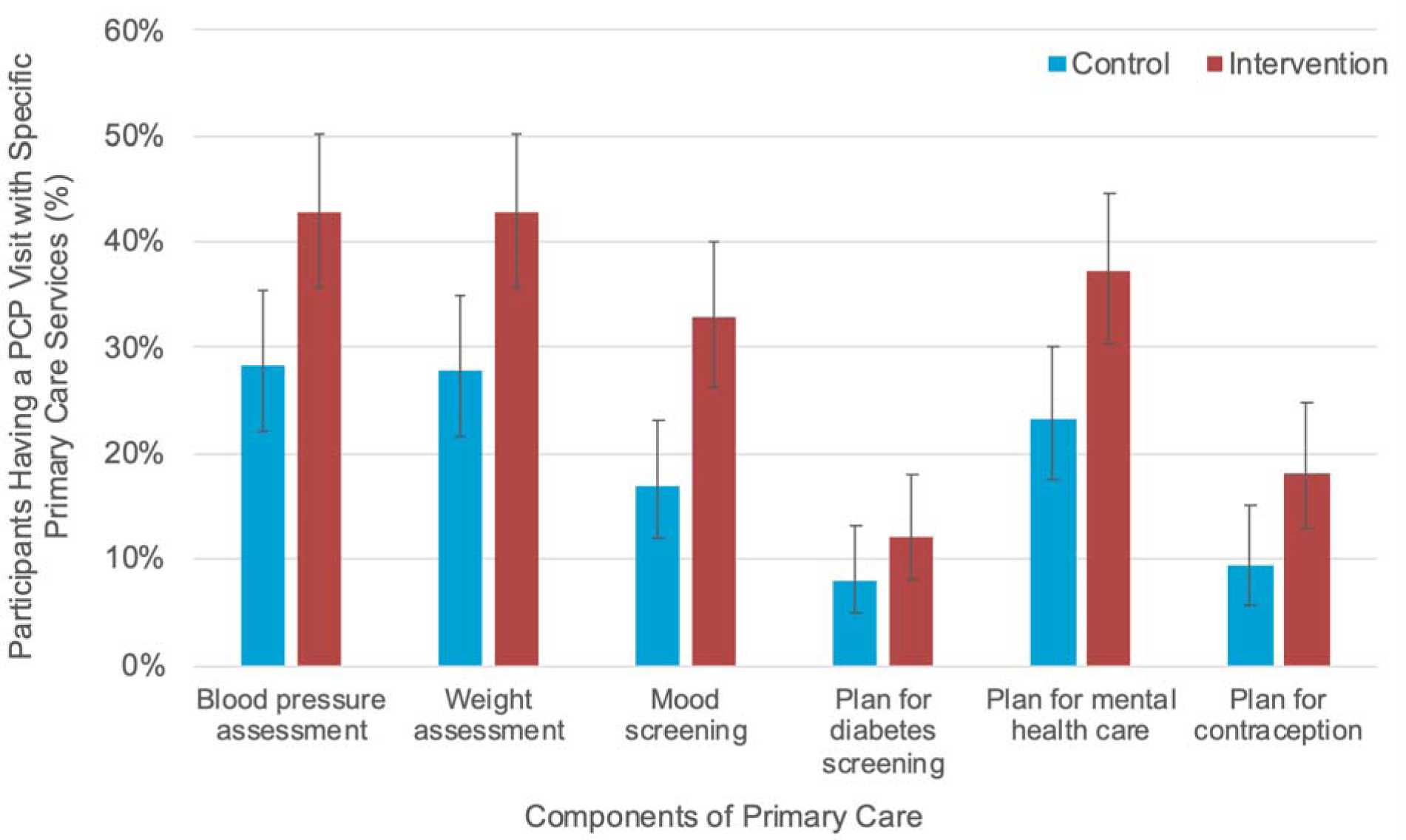
Effects on Content of Care Received by a Primary Care Practitioner. The figure presents the effects of the intervention on the percent of participants in each arm who had a primary care practitioner (PCP) visit that included the receipt of individual services (as listed in the horizontal axis) within 4 months postpartum. Outcomes are not contingent on having a primary care visit.

There was treatment effect heterogeneity across health conditions, demographic characteristics, and baseline self-reported physical and mental health (eTable 5). While the study was not powered to detect outcomes within subgroups, the intervention was associated with increases in PCP visits among nearly all subgroups examined.

## DISCUSSION

Among pregnant people with common comorbidities, a behavioral economics-informed intervention bundle, including default appointment scheduling, tailored messaging, and nudge reminders, increased PCP visit completion within 4 months postpartum by approximately 20 percentage points, a nearly 2-fold increase. The primary finding was robust to multiple definitions or variations of the primary outcome, including self-reported PCP visit attendance. The effects on the primary outcome appeared largely consistent among population subgroups, though small sample sizes limited power in these comparisons. Not only did the intervention increase PCP visit completion, but it also resulted in more individuals receiving important screening tests and services. There were no observed changes in emergent or urgent care visits between the two groups. However, any potential effects of facilitated primary care engagement on emergent care use are more likely to occur later in the postpartum year or beyond, and we intend to measure longer-term care utilization and outcomes in future studies.

Our results suggest that behavioral economic-informed interventions that reduce patient administrative burden have the potential to be relatively low-resource, high-impact approaches to increasing primary care use, a critical priority in the context of declining and inequitable primary care engagement in the U.S.^34,35^ Behavioral economics research examines how people make predictable decision errors and tests interventions that leverage these insights to remove behavioral barriers (“nudges”).^36–46^ These interventions often try to make it easier for people to make choices they already want to undertake but do not. In-kind, the underlying hypothesis of the present study was that many postpartum individuals with or at high risk for chronic conditions who have a PCP assigned want to be under the care of their PCP but face multiple barriers to primary care engagement in the postpartum period, including identifying who their PCP is and scheduling with them. Our study design was built to address two common behavioral barriers, namely, inattention and status-quo bias, and demonstrated how default primary care appointment scheduling, a salient label for the appointment, and tailored SMS messages and appointment reminders can increase postpartum primary care engagement. Similar approaches have effectively motivated other health behaviors, including in obstetric and postpartum care.^46–50^

This study builds on prior efforts to improve postpartum health and wellbeing.^51–58^ Our study is most closely aligned with the intervention research on postpartum care navigation, in which patient navigators identify and holistically address patient-level barriers to care and assist with care coordination.^51,59^ Although obstetric care navigators hold great promise for improving postpartum health care use, that level of intervention intensity and cost may not be necessary for most postpartum people needing primary care. Results from this study suggest that reducing some patient administrative barriers may be a relatively resource-conscious but highly effective approach to encouraging postpartum primary care transitions. Specifically, we demonstrated this intervention could be delivered consistently, with the successful scheduling of an “annual visit” appointment for 76% of participants and a low “no-show” appointment rate of only 4%. Future work should focus on examining the health impacts and cost-effectiveness of the intervention.

### Limitations

The study had several limitations. First, the study tested a bundled intervention; we were unable to measure the effectiveness of the individual components for increasing PCP visits. Next, we observed health care encounters within a single health system, though the health system is large (>1,300 PCPs). This study was also conducted in Massachusetts, in which pregnancy-related Medicaid coverage extends for 12 months postpartum and may impact the generalizability of our findings. We could not observe PCP visits for clinicians who do not use or are not affiliated with the health system’s common EHR (Epic). As an alternate measure, we did examine self-reports of PCP visits, which was highly consistent with results using EHR data. However, the response rate of approximately 62% (balanced across treatment and control groups) may also limit generalizability of self-reported outcomes. This study focused on individuals who had an identified PCP at enrollment; given the limited availability of PCPs in certain areas, the effect of the intervention may be lessened for individuals seeking to establish care with a new PCP. Last, the study was not powered to detect differences in many secondary outcomes related to the content of primary care within health conditions, and larger studies are needed to ascertain the impact of the intervention on the quality of primary care for specific conditions.

### Conclusions

In conclusion, a behavioral economics-informed intervention to improve postpartum transitions to primary care substantially increased postpartum primary care visit completion for patients with or at risk for common comorbidities. Targeting this population at a time of high health activation and motivation, this intervention represents a potentially scalable approach to increasing primary care engagement and ongoing health condition management in the postpartum months and beyond. Ongoing follow-up related to this study seeks to analyze the intervention’s impact on condition-specific management (i.e., the content and quality of care provided in the postpartum period) and its long-term impact on health outcomes. Similarly, as many individuals still did not attend a PCP appointment within 4 months, even with the assistance of this intervention, additional investigations should focus on identifying and addressing remaining barriers to transitioning to primary care after pregnancy.

## Data Availability

All data produced in the present study are available upon reasonable request to the authors.

## Disclosures

Dr. Clapp is a scientific medical advisor and has private equity for Delfina Care, which is not related and was not involved in this study. Dr. Ganguli received consulting fees from F-Prime for advising unrelated to this work.

## Presentations

Initial results from this study were presented at the University of Pennsylvania Center for Health Incentives and Behavioral Economics Research Conference (October 30, 2023; Philadelphia, PA), the 2023 Association for Public Policy Analysis and Management (APPAM) Fall Research Conference (November 9-11, 2023; Atlanta, GA) and the American Economic Association Annual Meeting (January 5-7, 2024; San Antonio, TZ). The study will be presented at the 2024 American Society for Health Economists conference (June 16-19, 2024; San Diego, CA).

## Previous Publication

A version of this manuscript was posted on the preprint server medrxiv.org (https://doi.org/10.1101/2024.01.21.24301585) so that it could be referenced in a recent grant proposal.

## Funding

This study was funded by the National Institute on Aging via the MIT Roybal Center for Translational Research to Improve Health Care for the Aging (P30AG064190) and the NBER Roybal Center for Behavior Change in Health (P30AG034532). Dr. Ganguli was supported by K23AG068240 from the National Institute on Aging. Additional support was provided by the National Academy of Medicine’s Health Catalyst Award. The funders had no role in the design and conduct of the study; collection, management, analysis, and interpretation of the data; preparation, review, or approval of the manuscript; or the decision to submit the manuscript for publication.

## Acknowledgments

The authors would like to thank Hasan Quadri and Fowsia Warsame for assisting in conducting this trial and Amanda Lee and Fatima Vakil from J-PAL North America for providing Research Management Support. Drs. Clapp and Cohen had full access to all the data in the study and take responsibility for the integrity of the data and the accuracy of the data analysis.

## Data Sharing Statement

Primary data and source code will be made publicly available at the Abdul Latif Jameel Poverty Action Lab Dataverse (https://dataverse.harvard.edu/dataverse/jpal) once planned secondary analyses have been completed (expected by August 2026).

## Notes

### Competing Interest Statement

The authors have declared no competing interest.

### Clinical Trial

NCT05543265

### Funding Statement

This study was funded by the National Institute for Aging via the MIT Roybal Center for Translational Research to Improve Health Care for the Aging (P30AG064190) and the NBER Roybal Center for Behavior Change in Health (P30AG034532). Additional support was provided by the National Academy of Medicine's Health Catalyst Award.

### Author Declarations

Ethics committee/IRB of Mass General Brigham gave ethical approval for this work.

### Summary of Updates

The manuscript and its title were updated after feedback from peer review. The analysis and results were not substantively changed.

