## Supplementary material for "Increasing Postpartum Primary Care Engagement through Default Scheduling and Tailored Messaging*: A Randomized Clinical Trial*": Online Supplement

### Supplement Contents


eFigure 1 ..... Example of Study Messages and Reminders

eTable 1 ...... Outcome Definition Table
eTable 2 ...... Comparison of the Intervention Components between Groups
eTable 3 ...... Reasons the Primary Care Appointment Could Not Be Scheduled
eTable 4 ...... Secondary Outcomes by Health Condition Subgroups
eTable 5 ...... Primary Outcome by Population Subgroups (Heterogeneity Analysis)


### eFigure 1. Examples of Study Messages and Reminders

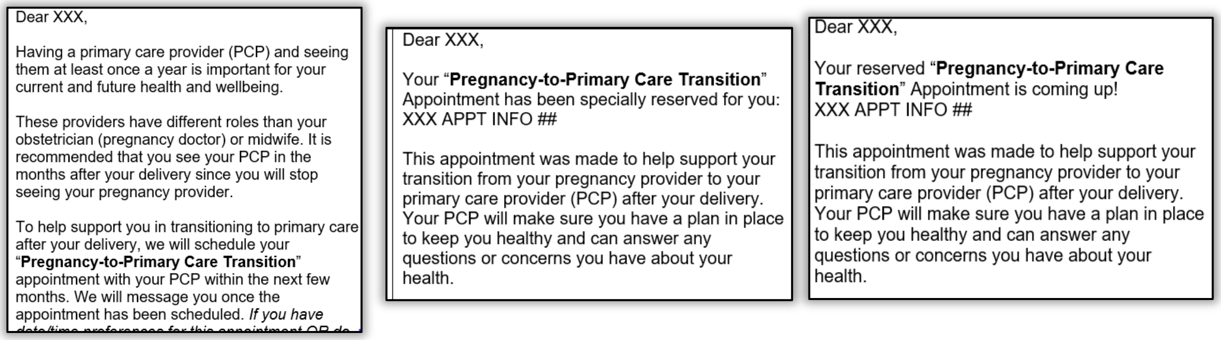

Dear XXX,

Having a primary care provider (PCP) and seeing them at least once a year is important for your current and future health and wellbeing.

These providers have different roles than your obstetrician (pregnancy doctor) or midwife. It is recommended that you see your PCP in the months after your delivery since you will stop seeing your pregnancy provider.

To help support you in transitioning to primary care after your delivery, we will schedule your "**Pregnancy-to-Primary Care Transition**" appointment with your PCP within the next few months. We will message you once the appointment has been scheduled. *If you have* [illegible]

Dear XXX,

Your "**Pregnancy-to-Primary Care Transition**" Appointment has been specially reserved for you:
XXX APPT INFO ##

This appointment was made to help support your transition from your pregnancy provider to your primary care provider (PCP) after your delivery. Your PCP will make sure you have a plan in place to keep you healthy and can answer any questions or concerns you have about your health.

Dear XXX,

Your reserved "**Pregnancy-to-Primary Care Transition**" Appointment is coming up!
XXX APPT INFO ##

This appointment was made to help support your transition from your pregnancy provider to your primary care provider (PCP) after your delivery. Your PCP will make sure you have a plan in place to keep you healthy and can answer any questions or concerns you have about your health.

**eTable 1. Outcome Definition Table**

Unless otherwise specified, all electronic health record (EHR)-based outcomes were assessed within 4 months of the estimated due date (EDD).

Self-reported outcomes occurred at the time of endline survey completion, unless otherwise noted. Endline surveys were sent to participants approximately 5 months after the EDD.

The trial protocol specified an a priori plan to examine outcomes among the different eligibility conditions (e.g., hypertension, diabetes). Those included in the clinicaltrials.gov registry (NCT 05543265) at the time of initial study entry (9/13/22) are designated in the table.

| Outcome | Source | Description | Pre-registered on clinicaltrials.gov |
|---|---|---|---|
| *Primary Outcome* | | | |
| Completion of a primary care visit for routine or chronic condition care within 4 months of EDD | EHR | Primary care practitioners are defined as those affiliated with family medicine, internal medicine, pediatrics, and gynecology practices.<br>Health care maintenance (or "annual exam") was determined by appointment "reason for visit," provider documentation, and diagnosis codes.<br>Condition-Specific Management was defined as a PCP visit for any reason in which one of the study's eligibility conditions were addressed in the provider's plan. | X |
| *Sensitivity Analysis Outcomes* | | | |
| Self-reported PCP Visit | Survey | Question: "When was the last time you saw a primary care provider for any reason? This does not include visits to your pregnancy care provider (OB/GYN or midwife), an emergency department, or urgent care center."<br>Positive Responses: "0-2 months after giving birth" or "3-4 months after giving birth" | X |
| Primary Outcome Restricted to Visits with Assigned PCP | EHR | Assigned PCP was the PCP listed in the patient's EHR as their PCP at the time of study enrollment. | X |
| Primary Outcome Restricted to Patients with PCP in the Same Health System | EHR | PCP in the Same Health System was defined as a PCP affiliated with the study institution's health system. | |
| Any Primary Care Visit within 4 months of EDD | EHR | Any primary care visit was defined as a visit with a physician or advanced practice provider affiliated with a family medicine, internal medicine, pediatrics, or gynecology clinic for any reason. | |
| Any Primary Care Visit or Scheduled within 1 Year of EDD | EHR | A scheduled visit was an upcoming visit that was scheduled but had not yet occurred at the time of chart review for the primary outcome. | X |
| *Secondary Outcomes in Overall Sample* | | | |
| Obstetric Triage Visit | EHR | Obstetric triage visit was defined as a presentation to an L&D triage unit for evaluation within 4 months of the EDD. | |
| Any Emergency Department or Urgent Care Visit | EHR | Emergency and urgent care were defined by visits occurring at those designated locations within the health system within 4 months of the EDD. | X |
| Postpartum Readmission | EHR | Postpartum readmission was defined as any readmission within 4 months of the EDD. | |

| | | | |
|---|---|---|---|
| Weight Assessment by PCP | EHR | Weight assessment was defined as a weight logged in the EHR at the PCP visit. | |
| Blood Pressure Assessment by PCP | EHR | Blood pressure assessment was defined as a blood pressure logged in the EHR at the PCP visit. | |
| Mood Screening by PCP | EHR | Mood screening was considered to be the use of any validated mood screening tool by the PCP. | |
| PCP Plan for Glucose Screening | EHR | Plan for glucose screening was considered any documentation of a discussion regarding diabetes/glucose screening, an order for a screening test, or inclusion of a recent glucose screening test result within a PCP visit note. | |
| PCP Plan for Mental Health | EHR | Plan for mental health care was considered any documentation of a discussion regarding mental health (namely, anxiety and depression) in a PCP visit note. | |
| PCP Plan for Contraception | EHR | Plan for contraception was considered any documentation of a discussion regarding contraception, regardless of use or type, within a PCP visit note. | X |
| *Secondary Outcomes by Subgroups of Health Conditions* | | | |
| Weight Assessment by PCP among those with Pre-pregnancy Obesity | EHR | Weight assessment was defined as a weight logged in the EHR at the PCP visit. | |
| PCP Documentation of a Plan for Weight Management among those with Pre-pregnancy Obesity | EHR | Weight assessment was defined as a weight logged in the EHR at the PCP visit. | X |
| Mental Health Service Referral or Use among those with Anxiety or Depression | EHR | Mental health services were considered to be care provided by social workers, therapists, psychologists, and psychiatrists. | X |
| Medication Use for Mood or Anxiety Disorder among those with Anxiety or Depression | EHR | Medication use was ascertained from the patient's medication list and from the clinical documentation. | X |
| Mood Screening by PCP among those with Anxiety or Depression | EHR | Mood screening was considered to be the use of any validated mood screening tool by the PCP. | |
| PCP Plan for Mental Health among those with Anxiety or Depression | EHR | Plan for mental health care was considered any documentation of a discussion regarding mental health (namely, anxiety and depression) in a PCP visit note. | |
| Blood Pressure Assessment by PCP among those with a Chronic or Pregnancy-related Hypertensive Disorder | EHR | Blood pressure assessment was defined as a blood pressure logged in the EHR at the PCP visit. | X |
| Antihypertensive Medication Use among those with a Chronic or Pregnancy-related Hypertensive Disorder | EHR | Medication use was ascertained from the patient's medication list and from the clinical documentation. | X |
| Diabetes Screening Test Among Individuals with Gestational Diabetes | EHR | Diabetes screening tests were defined as Hemoglobin A1c or Glucose Tolerance Test. | X |
| Glucose Control Assessment Among | EHR | Glucose control assessment was defined as Hemoglobin A1c. | X |

| | | | |
|---|---|---|---|
| Individuals with Pre-Gestational Diabetes | | | |
| Diabetes Medication Use Among Individuals with Pre-Gestational Diabetes | EHR | Medication use was ascertained from the patient's medication list and from the clinical documentation. | X |
| PCP Plan for Glucose Screening Among Individuals with Diabetes | EHR | Plan for glucose screening was considered any documentation of a discussion regarding diabetes/glucose screening, an order for a screening test, or inclusion of a recent glucose screening test result within a PCP visit note. | |

*PCP, primary care practitioner.*

### eTable 2. Comparison of the Intervention Components between Groups

| Intervention Component | Control (n=173) | Treatment (n=180) |
|---|---|---|
| Generic PCP Visit Recommendation Message Sent | 173 (100%) | - |
| Tailored PCP Visit Recommendation Message Sent | - | 180 (100%) |
| PCP Appointment Scheduled by Study Staff | - | 137 (76%) |
| PCP Appointment Scheduled within 4 months EDD | - | 124/137 (91%) |
| PCP Appointment but Patient did not Attend or Cancel (“no-show”) | - | 6/137 (4%) |
| Reminder PCP Appointment Messages Sent via EHR Patient Portal | - | 180 (100%) |
| Text Message (SMS) PCP Appointment Reminders Sent | - | 180 (100%) |
| Electronic Message Sent to PCP about Scheduled Appointment | - | 88 (49%) |

*Abbreviation: EDD, Estimated Due Date; EHR, Electronic Health Record; PCP, Primary Care Practitioner.*

### eTable 3. Reasons the Primary Care Appointment Could Not Be Scheduled

| Reason | Frequency n=43 |
|---|---|
| Patient had already had a follow-up scheduled in postpartum period | 21 (49%) |
| Patient needed to re-establish care or reported wanting to establish care with a new PCP | 10 (23%) |
| Patient had recent annual visit within 1 year | 5 (12%) |
| Study staff unable to contact PCP office or PCP office didn't allow study staff to make appointment | 2 (5%) |
| PCP office reported no record of patient or prior care | 2 (5%) |
| Insurance information not updated at PCP office | 2 (5%) |
| Patient placed on waitlist for visit | 1 (2%) |

### eTable 4. Secondary Outcomes by Health Condition Subgroups

| **Outcomes by Health Conditions** | **Control Group** n (%) | **Intervention Group** n (%) |
|---|---|---|
| Individuals with Obesity | | |
| PCP visit with weight assessment | 22/75 (29.0%) | 31/69 (45.0%) |
| PCP visit with documentation of a weight management plan | 9/75 (12.0%) | 15/69 (22.0%) |
| Individuals with Mental Health Condition | | |
| PCP visit including a referral for mental health services | 35/128 (27.3%) | 34/138 (24.6%) |
| Medication use for mood or anxiety disorder | 60/128 (46.9%) | 57/138 (41.3%) |
| PCP visit with mood screening | 20/128 (15.6%) | 46/138 (33.3%) |
| PCP visit with documentation of a mental health plan | 31/128 (24.2%) | 55/138 (39.9%) |
| Individuals with Hypertensive Disorder | | |
| PCP visit with blood pressure screen | 9/26 (35.0%) | 14/31 (45.0%) |
| Antihypertensive medication use | 9/26 (35.0%) | 8/31 (26.0%) |
| Individuals with Diabetes | | |
| Diabetes screening test among individuals with gestational diabetes | 15/28 (54.0%) | 20/27 (74.0%) |
| Glucose control assessment among individuals with pre-gestational diabetes | 3/10 (30.0%) | 1/4 (25.0%) |
| Diabetes medication use among individuals with pre-gestational diabetes | 7/10 (70.0%) | 3/4 (75.0%) |
| PCP visit with documentation of a plan for diabetes screening and/or management | 8/38 (21.0%) | 16/31 (52.0%) |

**eTable 5. Primary Outcome by Population Subgroups (Heterogeneity Analysis)**

| Subgroup | Control Group n (%) | Intervention Group n (%) | Adjusted Between-group difference, percentage point (95 CI) [5] |
|---|---|---|---|
| Site of Prenatal Care | | | |
| Hospital-based Clinic | 29/121 (24.0) | 49/129 (38.0) | 14.6 (3.1 to 26.1) |
| Community-based Clinic | 9/52 (17.3) | 23/51 (45.1) | 28.5 (11.1 to 46.0) |
| Health Condition | | | |
| Diabetes [1] | 9/38 (23.7) | 21/31 (67.7) | 46.3 (24.3 to 68.3) |
| Hypertension [1] | 7/26 (26.9) | 16/31 (51.6) | 24.2 (-0.2 to 50.8) |
| Mental Health Condition | 28/128 (21.9) | 56/138 (40.6) | 18.9 (7.9 to 30.0) |
| Obesity | 19/75 (25.3) | 27/69 (39.1) | 15.6 (0.2 to 31.1) |
| Multi-Morbidity [1] | 20/73 (27.4) | 33/67 (49.3) | 22.3 (6.3 to 38.4) |
| Race [2] | | | |
| Asian | 3/14 (21.4) | 3/11 (27.3) | 3.6 (-32.6 to 39.8) |
| Black | 5/14 (35.7) | 8/14 (57.1) | 22.6 (-16.0 to 61.2) |
| Multiple Races or “Other” [3] | 5/26 (19.2) | 11/25 (44.0) | 29.7 (2.5 to 57.0) |
| White | 25/117 (21.4) | 50/127 (39.4) | 18.4 (6.8 to 30.0) |
| Ethnicity [2] | | | |
| Non-Hispanic | 29/127 (22.8) | 57/139 (41.0) | 18.8 (7.7 to 29.9) |
| Hispanic | 6/41 (14.6) | 14/37 (37.8) | 23.7 (3.9 to 43.5) |
| Individual Annual Earnings [2] | | | |
| <$30,000 | 8/32 (25.0) | 17/36 (47.2) | 21.0 (-2.1 to 44.0) |
| $30,000-74,999 | 8/37 (21.6) | 23/55 (41.8) | 20.6 (-0.3 to 40.6) |
| ≥$75,000 | 16/82 (19.5) | 29/76 (38.2) | 19.4 (5.0 to 33.6) |
| Primary Payer for Delivery Hospitalization | | | |
| Medicaid | 9/40 (22.5) | 14/35 (40.0) | 17.8 (-3.4 to 29.1) |
| Private / Other | 26/130 (20.0) | 53/138 (38.4) | 18.9 (8.1 to 29.7) |
| Self-Reported Physical Health [4] | | | |
| “Good”, "Very Good," or "Excellent" | 31/151 (20.5) | 60/161 (37.3) | 17.4 (7.4 to 27.4) |
| “Fair” or “Poor” | 7/22 (31.8) | 12/19 (63.2) | 27.4 (-5.2 to 59.9) |
| Self-Reported Mental Health [4] | | | |
| “Good”, "Very Good," or "Excellent" | 32/136 (23.5) | 57/148 (38.5) | 15.4 (4.6 to 26.2) |
| “Fair” or “Poor” | 6/37 (16.2) | 15/32 (46.9) | 31.4 (10.2 to 52.6) |

[1] Includes both pre-pregnancy and pregnancy-related conditions.
[2] Self-reported.
[3] Patients could select “Other” as a race option if they did not self-identify with the other options: American Indian or Alaskan Native, Asian, Black, Native Hawaiian or Other Pacific Islander, White).
[4] Self-reported at time of enrollment.
[5] To account for randomization strata, regressions include indicator variables for whether or not the participant had any primary care practitioner (PCP) visit in the 3 years before randomization and whether the participant received prenatal care from a hospital or health center.
